# Development and accuracy of a novel machine learning model to detect toddlers’ physical activity and sedentary time using accelerometers: Little Movers Activity Analysis

**DOI:** 10.1101/2025.04.24.25326266

**Authors:** Elyse Letts, Sara King-Dowling, Natascja Di Cristofaro, Patricia Tucker, John Cairney, Dylan Kobsar, Brian W. Timmons, Joyce Obeid

## Abstract

**Objective:** (1) develop and test a novel, open-source, supervised machine learning model to detect toddlers’ physical activity (PA) and sedentary time (SED); (2) compare this novel machine learning model to existing cut-point methods to analyse toddlers’ PA (independent sample cross-validation of existing methods). **Methods:** We recruited 111 healthy toddlers to attend two semi-structured lab visits while wearing an ActiGraph wGT3X-BT accelerometer. Sessions were video recorded and manually annotated using a modified Children’s Activity Rating Scale to determine a ground truth of toddler activity. We extracted 40 time and frequency domain features from the raw accelerations (across 6 different epochs ranging from 1-60s) and trained 4 gradient boosted tree machine learning models. Models were assessed using accuracy, F1 scores, and confusion matrices. For the validation of existing methods, we calculated accuracy, F1, and mean absolute differences (MAD) in total PA (TPA) and moderate-to-vigorous PA (MVPA) estimation. **Results:** The 5s epoch performed best with the recommended models classifying non-volitional movement (NVM)/SED/TPA and NVM/SED/light PA (LPA)/MVPA reaching an 82% and 74% accuracy with MAD of 3.0 and 3.2min/hour, respectively. Independent sample cross-validation found accuracies from 33-73% and MAD ranging from 7.6-18.6min/hour in TPA and 10.7-25.8min/hour in MVPA. **Conclusions:** We recommend the NVM/SED/TPA or NVM/SED/LPA/MVPA models given alignment with toddler TPA guidelines and MVPA link to health outcomes, respectively. We additionally present an open-access, user-friendly interface for using these models that does not require coding knowledge. This study presents a substantial step forward toward comprehensive and accessible measurement of toddlers’ physical activity.

**Summary Box:**

- **What is already known on this topic** - Current methods used to assess toddlers’ physical activity (PA) and sedentary time (SED) face challenges with low accuracies and inabilities to detect non-volitional movement (NVM, e.g., being carried). Machine learning methods are promising but existing methods have been validated using small samples and the algorithms are not openly available.
- **What this study adds** – Open-source machine learning models to detect toddlers’ PA, SED, and NVM that outperform existing available cut-point methods, including an easy-to-use interface to apply these models to raw accelerometer data.
- **How this study might affect research, practice or policy** – We provide an easy-to-use, open access tool for researchers, clinicians, and beyond to use machine learning to more accurately detect toddlers’ volitional PA and SED. This will allow for more accurate quantifications of PA that can be linked to health outcomes and inform toddler PA guidelines.

## Introduction

Current guidelines recommend toddlers (ages 1-2 years) engage in 180 minutes of daily physical activity [1–3]. Adherence to these guidelines has varied widely (0-98%) [4–6], partly due to differences in measurement (e.g., questionnaires, different accelerometer cut-points [7]). Parent-reported data lack reliability, especially with multiple caregivers involved [8]; accelerometers offer promise but existing validation studies have consistently yielded lower accuracy [4].

A 2020 review identified 10 accelerometer cut-point methods to assess toddlers’ physical activity (PA) [4] with the most common from Trost et al., 2012 [9]. These methods, developed for toddlers or older children, struggle to detect non-volitional movement (NVM; e.g., being carried, which occupies a significant part of a toddler’s day) and may miss key behaviors like crawling or unstable gait. Count-based metrics reduce activity variability and are device-specific [10].

Machine learning (ML) is a promising option to more accurately capture toddler activity using raw accelerations. While its use has rapidly expanded in adult accelerometry analysis [11–14], few methods have been tested with toddlers [15]. To our knowledge, only five toddler-specific models have been developed, four of which are supervised learning models (trained on a ground truth of direct observation). These are a support vector machine [16] and random forest models [17–19], with accuracies from 64% - 89%. However, the sample sizes are small (10-24 participants) with limited activities due to the short (6-25 minutes/participant) observation periods [17,19]. The unsupervised model is a hidden semi-Markov model [20] using clustering of data of 279 children aged 9-38 months. While promising, it has no ground truth validation, meaning its true performance is unclear. Further, none of these models are publicly available which limits their use [21]. Supplementary Material 1 provides details on these models.

This study aimed to fill these gaps by developing a novel, open-source, supervised ML model to detect PA, sedentary time (SED), and NVM in a large sample of toddlers and assess its accuracy against direct observation (Phase I and II validation [22]). A second objective was to compare this ML model to existing available methods, providing an independent sample cross-validation (Phase III validation [22]).

## Methods

This cross-sectional study recruited 111 toddlers as part of the **i**nvestigating the validity and reliability of accelerometer-based measures of **P**hysica**L A**ctivity and sedentar**Y** time in toddlers (iPLAY) study. Toddlers were recruited from the Hamilton community in Ontario, Canada through childcare centres, community events, websites, and social media. Participants were 12.0-35.9 months of age with no known physical disability or medical condition that impacted movement. This study received ethics clearance from the Hamilton integrated Research Ethics Board (HiREB #3674). Informed written consent was obtained from a parent of each participant. Data were collected from February 2019 through March 2020.

### Data collection

Eligible participants attended two, ∼1-hour lab visits separated by ∼1 week. Participants wore an ActiGraph wGT3X-BT accelerometer on an elastic belt around the waist, over the right hip (30 Hz sampling frequency, dynamic range ± 8*g*), and were video recorded using two cameras (Sony HDR-AS50). The right hip was chosen as a more feasible wear location than wrist for toddlers [23], reflecting whole body movement and energy expenditure [24]. The video and accelerometer were synched to the millisecond by displaying the laptop (used to initialize accelerometer) time during each video. The research team conducted a gross motor skill assessment (Peabody Developmental Motor Scales – 2^nd^ ed [25]) and guided participants through eight stations of toddler activity interspersed with free-play. Stations included: laying down, sitting to read, sitting to draw, floor-based activities (e.g., playing with blocks), walking (with assistance if needed), jumping, running, stroller/carried. Parents completed a demographic/medical questionnaire.

### Ground truth video annotation

Following the visits, all videos (n = 218; average length: 64.10 minutes; minimum/maximum: 43.48/100.10 minutes) were uploaded to a custom software [26] and manually annotated second-by-second to label the ground truth of the toddler activity. For videos with two participants in the same session (e.g., siblings), videos were coded twice, once for each child. Direct observation was chosen as the ground truth to best capture the context and variation in toddler activity (e.g., touching he accelerometer, NVM) [11]. Annotations were a modified version of the Children’s Activity Rating Scale (CARS), which has been validated against energy expenditure in young children [27]. Modifications added other activity codes (e.g., touching accelerometer, NVM) and position (sit/stand/lie/other). Table 1 shows the complete annotation scheme (detailed annotation instructions are in Supplementary Material 2).

**Table 1.**
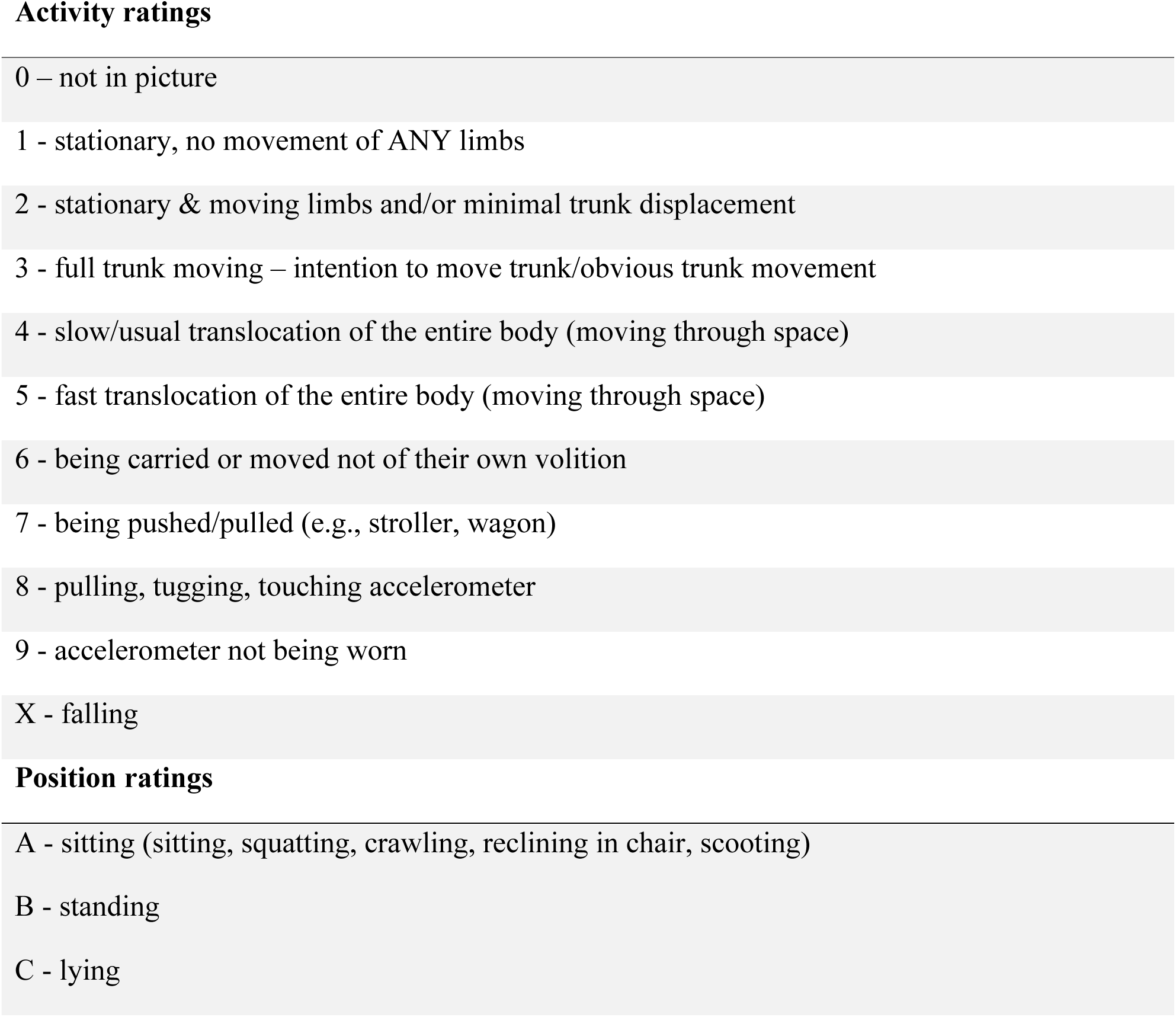
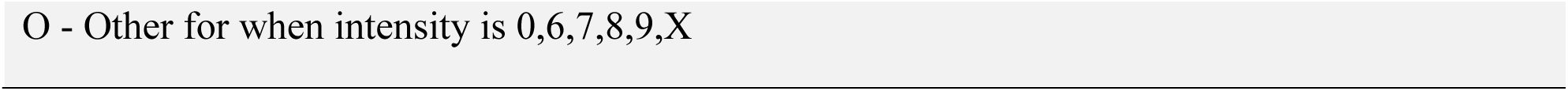
Video annotation scheme for direct observation annotation.

Annotation was completed by 19 raters who underwent training on six 5-minute clips, having to reach at least 80% agreement with the expert rater (EL). During the annotation process, 22 full videos (10% of videos) were randomly selected to be annotated by two raters for reliability analyses (15 raters). Activity ratings had an intraclass correlation of 0.83, considered good [28], and a Fleiss’ Kappa of 0.61, considered substantial agreement [29]. Position ratings had a Fleiss’ Kappa of 0.85, considered near perfect agreement, and a Krippendorff’s alpha of 0.85, considered acceptable (>0.67) [30]. When adapted for epochs longer than 1 second, the highest mode was used for activity and transitions in position were maintained (e.g., 3 seconds of 3B,3B,2A would become 3BA for a 3-second epoch). These annotations were converted to intensity classifications as follows: NVM = 6, 7; SED = 1, 2(sitting or lying); light PA (LPA) = 2(standing), 3; moderate-to-vigorous PA (MVPA) = 4, 5; total PA (TPA) = 2(standing), 3, 4, 5. This aligns with previous observations of energy expenditure in preschool children [31,32] and with the original CARS codes [27].

### Accelerometer processing

Raw acceleration .gt3x files were downloaded and converted to .csv files using pygt3x (v0.5.2) in python (v3.9.6). We then extracted 40 time and frequency features using the python tsfresh package (v0.20.1) for the x, y, z, and vector magnitude of the raw accelerations: mean, standard deviation, minimum, maximum, quantiles (10, 25, 50, 75, 95), coefficient of variation, sum of values, median, skewness and kurtosis (using adjusted Fisher-Pearson standardized moment coefficient), root mean square, fast Fourier transformation (FFT) entropy (binned entropy of the power spectral density of the time series, using the welch method, and 10 bins), aggregated FFT (spectral centroid (mean), variance, skew, and kurtosis of the absolute Fourier transform spectrum), and the real, imagined, absolute, and angle of the first 4 coefficients from the FFT. Features were selected from previous literature and limited to 40 to balance scope and computation requirements [16,17,33]. All features were extracted for non-overlapping 1s, 3s, 5s, 15s, 30s, and 60s epochs.

### Machine learning model development

Data were divided into training, cross-validation, and testing sets using a 60/20/20% ratio. The sets were divided by participant ensuring no data leakage, meaning that all of one participant’s data (both visits) were only in one set. The training set contains data from 66 participants across 129 visits. The ML model used is gradient-boosted trees classification using XGBoost (v2.0.0) and sklearn (v1.2.2) in python (v3.9.6). A gradient-boosted trees model is a decision tree ensembling method (similar to a random forest model, which showed the highest accuracy in other toddler models [18]) which creates sequential trees that learn from each other [34]. It is generally considered state-of-the-art for classification tasks such as this one [35,36].

Hyperparameters were tuned on the cross-validation set using a sequential model-based optimization using gbrt_minimize (Gradient Boosted Regression Tree surrogate model) from scikit-optimize (skopt v0.10.2) with 20 random starts and 70 evaluations. Parameters tuned were learning_rate, n_estimators (number of trees), max_depth (tree maximum depth), min_child_weight (the minimum weight needed in a child node), gamma (minimum loss reduction to make a split), subsample (proportion of samples used for each tree), and colsample_bytree (the proportion of features used for each tree). Sample weights were manually tuned to better account for class imbalances in outcomes.

Four final models were trained with four sets of outcomes based on common research outcomes and adding NVM: (1) SED/TPA; (2) SED/LPA/MVPA; (3) NVM/SED/TPA; and (4) NVM/SED/LPA/MVPA. For all models, windows with activity codes of 0, 8, 9, and X indicating missing or invalid data were removed prior to training. For models 1 and 2, windows classified as NVM (codes 6 and 7) were also removed. Each model was evaluated using the accuracy, F1 score, and confusion matrices. F1 scores are the harmonic mean of precision and recall (considering true positives, false positives, and false negatives) [37] and are better able to account for class imbalances in the data for which accuracy may provide an overconfident estimate. Confusion matrices are used to understand directions of misclassification and class level accuracy. Previous studies on XGBoost models indicate that a minimum sample size of 20 [38] is required to have sufficient power to detect accuracies greater than 95%, so our sample size of 111 (66 in the training set) greatly exceeds this minimum.

### Independent cross sample validation of existing methods

To address the second objective, we tested 11 openly-available methods that have been used in the toddler population, details of which are shown in Table 2. These waist-worn methods were identified from previous reviews and studies [4,9]. We applied the methods to our data and compared to our ground truth video annotations. We calculated each method’s accuracy and F1 score using the full 111 participants across all visits, with NVM removed as no methods available in the literature detect NVM. We also compared the time in TPA and MVPA calculated by each method with ground truth (mean absolute and relative differences) to understand the extent to which methods may under/overestimate PA. For TPA estimation, Pulakka 2013 and Pate 2006 methods were removed as they combine SED and LPA so cannot estimate TPA. For MVPA estimation, the Dorris 2025 and Oftedal 2014 methods were removed as they only assess TPA, not MVPA.

**Table 2.**
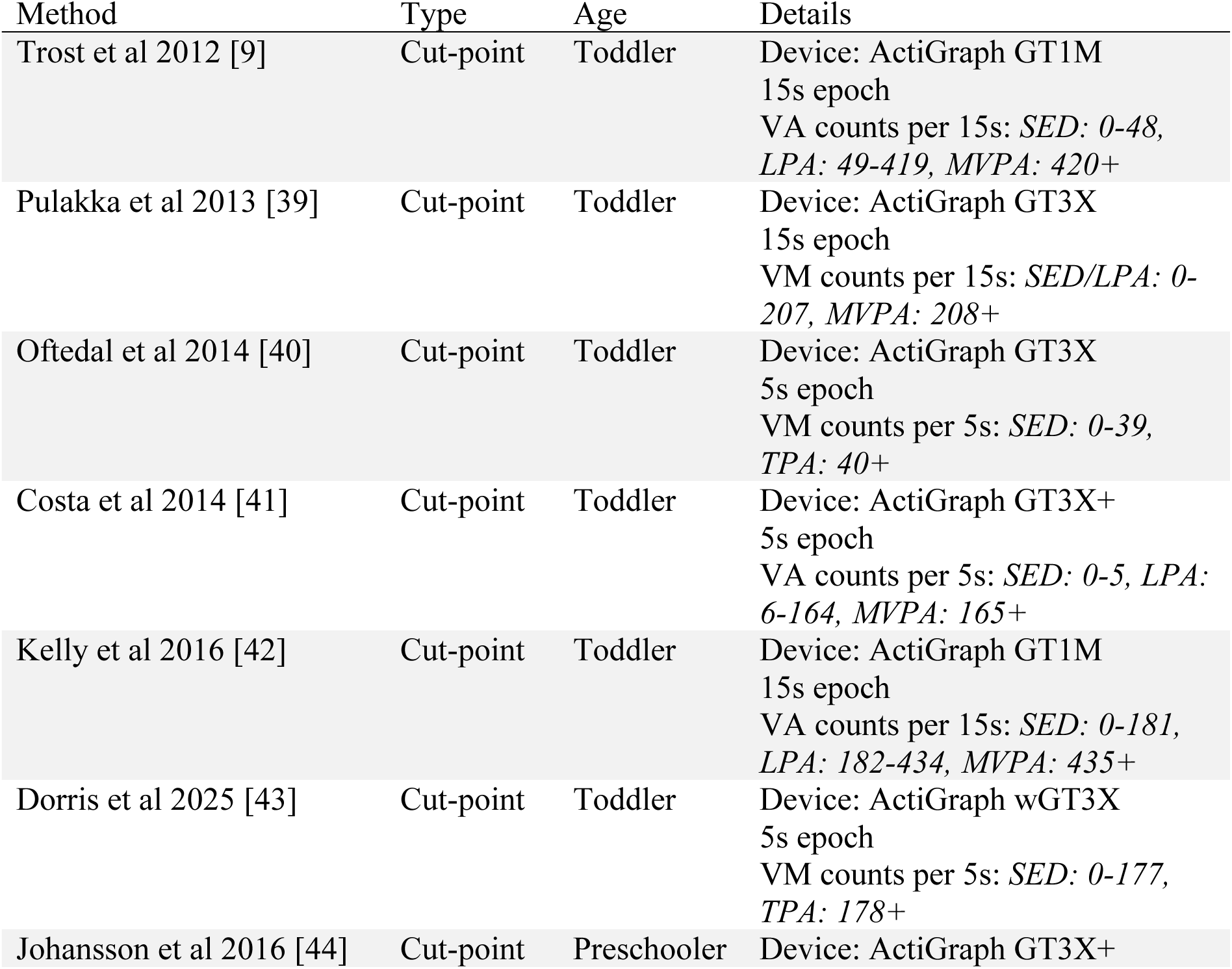
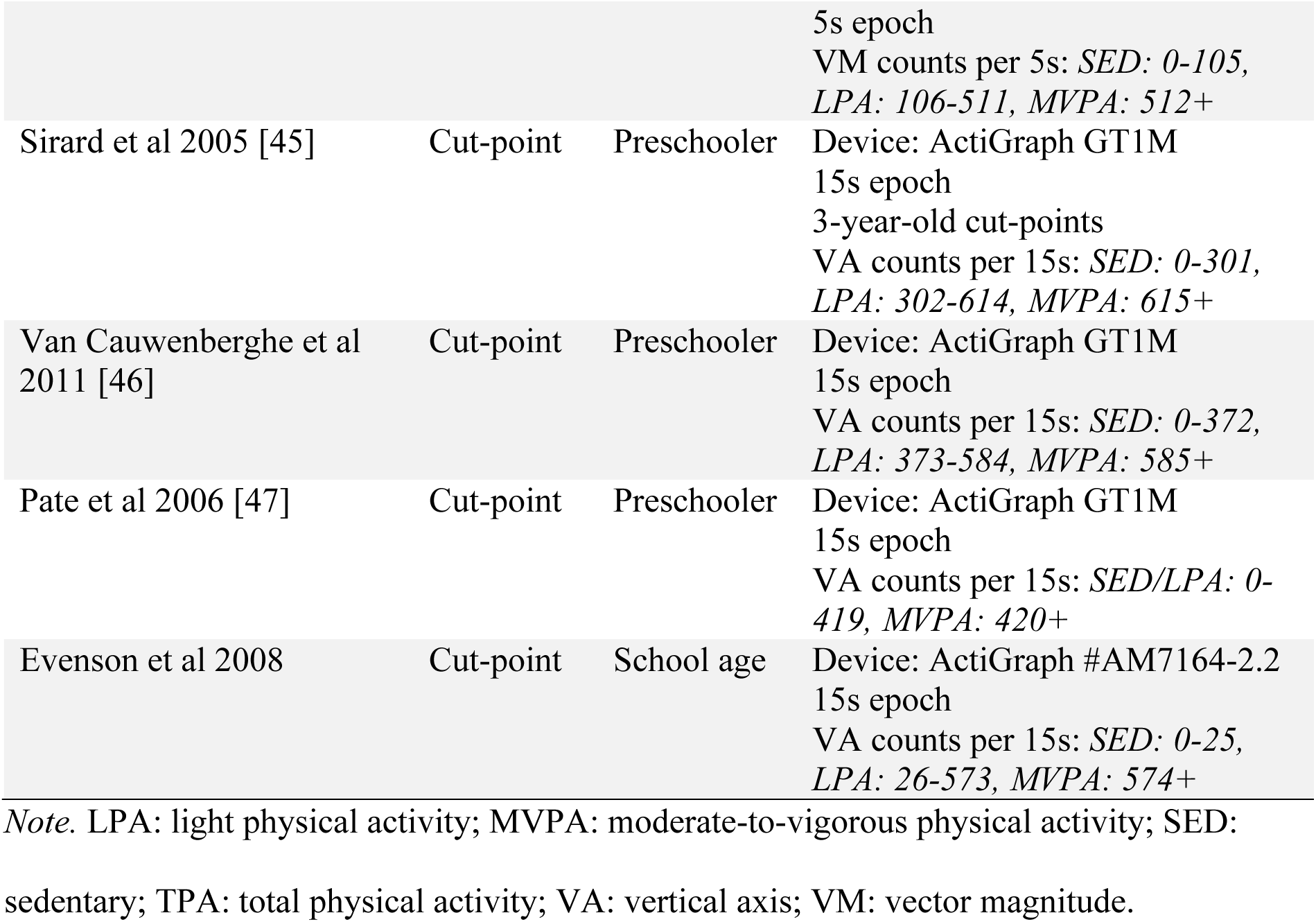
Methods tested in the independent sample cross-validation.

### Patient and Public Involvement statement

Patients or the public were not involved in the design, conduct, reporting, or dissemination plans of our research.

### EDI statement

Participant recruitment was designed to be as reflective of the Hamilton, Canada community by advertising through child-care centres, flyers, websites, and social media. Inclusion was limited to healthy toddlers for initial model development, but further validation in toddlers with chronic medical conditions and disabilities will be the next phase of our study. Full participant characteristics are available in Table 3, with an even split between males/females. The authors of this paper encompass different genders and career stages.

**Table 3.** Participant demographic details.

|  | Overall<br>(N = 111) |
| --- | --- |
| **Age (m)** |  |
| Mean (SD) | 21.3 (7.0) |
| Median [Min, Max] | 19.4 [12.0, 36.2] |
| **Sex** |  |
| Female | 57 (51%) |
| Male | 54 (49%) |
| **Prematurity** |  |
| Not premature | 94 (85%) |
| Premature ($\geq 3$ weeks) | 14 (13%) |
| Not reported | 3 (3%) |
| **Single/Multiple** |  |
| Single | 102 (92%) |
| Twin | 9 (8%) |
| **APGAR concern at birth** |  |
| No | 100 (90%) |
| Yes | 6 (5%) |
| Unsure | 5 (5%) |
| **Older sibling living in the home** |  |
| No | 62 (56%) |
| Yes | 49 (44%) |
| **Race/Ethnicity** |  |
| White | 80 (72%) |
| Multiracial | 23 (21%) |
| Asian | 5 (5%) |
| Latin American | 2 (2%) |
| Arab | 1 (1%) |
| **Home language** |  |
| English | 100 (90%) |
| Other<br>(Reported: Mandarin, Punjabi, Spanish, Portuguese, Arabic, Khmer,<br>Italian) | 8 (7%) |
| Not reported | 3 (3%) |
| **Parent 1 education** |  |
| College/university | 105 (95%) |
| High school | 4 (4%) |
| Not reported | 2 (2%) |
| **Parent 2 education** |  |
| College/university | 97 (87%) |
| High school | 7 (6%) |
| Not reported/not applicable | 7 (6%) |
| **Marital status** |  |
| Married | 88 (79%) |
| Living common-law OR living with a partner | 16 (14%) |
| Single, never married | 4 (4%) |
| Separated | 1 (1%) |
| Not reported | 2 (2%) |
| **Total annual household income before taxes** |  |
| Less than \$35,000 | 10 (9%) |
| Between \$35,000 and \$49,999 | 8 (7%) |
| Between \$50,000 and \$74,999 | 11 (10%) |
| Between \$75,000 and \$99,999 | 21 (19%) |
| Between \$100,000 and \$124,999 | 23 (21%) |
| Greater than \$125,000 | 36 (32%) |
| Prefer not to answer | 2 (2%) |

*Note.* SD = standard deviation. APGAR concern at birth refers to any concerns identified by a clinician regarding appearance (skin colour), pulse, reflexes, muscle tone, and respiration at birth.

## Results

### Participants

All 111 participants completed visit 1 and 108 completed both visits (unable to schedule visit 2 due to COVID-19 pandemic closures). One participant had an accelerometer malfunction for the second visit, resulting in 218 total visits included. Participants had a mean age of 21.3 months and 51% were female. Table 3 provides complete participant demographic information.

### Machine learning model results

When comparing epochs, we saw increases in accuracy from 1s-5s that stabilized from 15s-60s. F1 scores showed a similar trend, increasing up to 5s, then decreasing from 15s-60s. From the confusion matrices, the 5s epoch best captured NVM while maintaining the highest accuracy and F1 score (within 1%). As such, we used the 5s epoch for all remaining analyses. Supplementary Material 3 shows the full results and confusion matrices of epoch analyses. Supplementary Material 4 shows the frequency of each code and category from video annotations in the 5s epoch. From this, we can see a clear class imbalance where, for example, NVM has about 8x fewer observations than SED.

The four final models had accuracy and F1 scores as follows (accuracy%/F1%): (1) SED/TPA: 87/87; (2) SED/LPA/MVPA: 76/75; (3) NVM/SED/TPA: 82/69; and (4) NVM/SED/LPA/MVPA: 74/65. Confusion matrices for each are shown in Figure 1. NVM was consistently the most misclassified, followed by LPA. Supplementary Material 5 contains feature importance rankings.

**Figure 1.**
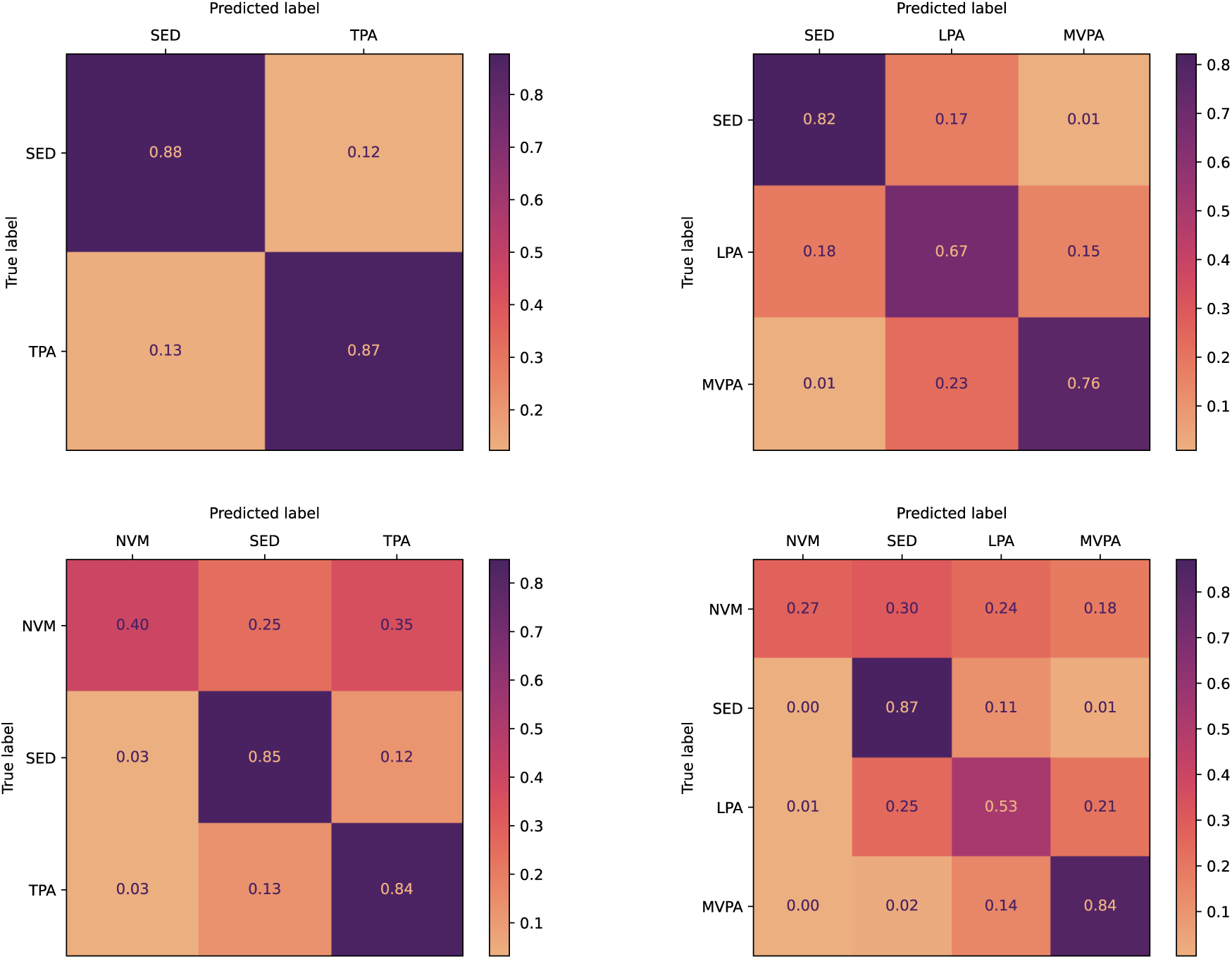
Confusion matrices for newly developed machine learning models. LPA: light physical activity; MVPA: moderate-to-vigorous physical activity; NVM: Non-volitional movement; SED: sedentary; TPA: total physical activity.

### Independent sample cross-validation

We calculated the accuracy and F1 scores for each of the 11 existing methods (Table 2), which ranged from 30-73%. Figure 2 (Panel A) shows the full details of accuracy and F1 scores for each method compared to the ML models.

**Figure 2.**
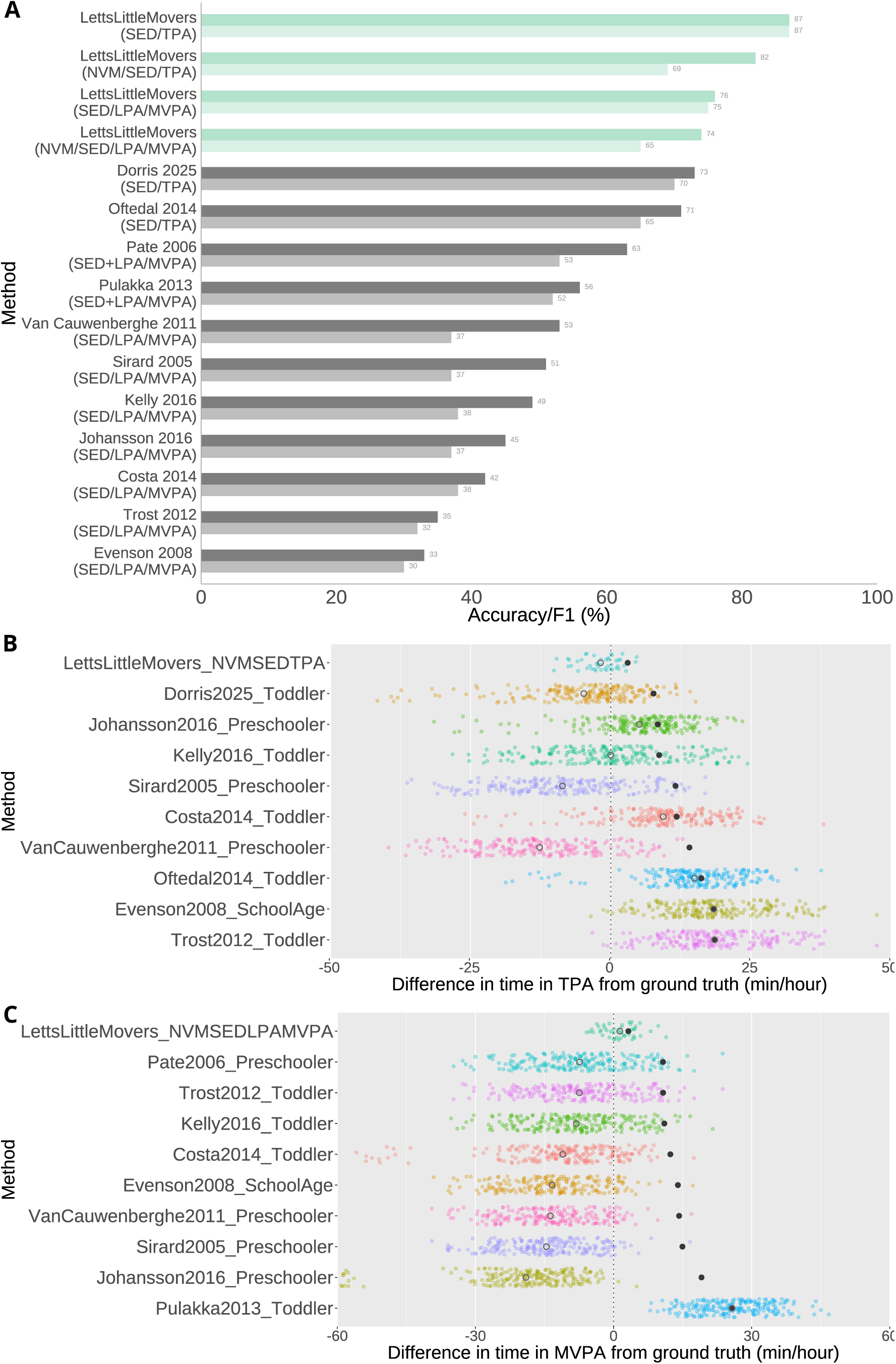
Results of independent sample cross validation compared to machine learning models. *Panel A:* Accuracy and F1 scores for the newly developed machine learning models (in green) and existing cut-point methods (in grey) used in toddler populations. Dark colour shows accuracy and light colour shows F1 score. *Panel B/C:* Difference in time in TPA (Panel B) and MVPA (Panel C) for each method compared to ground truth. Values are presented in minutes per hour. Solid black circles represent the mean absolute differences, open grey circles represent the mean relative differences, and coloured points show the relative differences for each participant visit. LPA: light physical activity; MVPA: moderate-to-vigorous physical activity; NVM: non-volitional movement; SED: sedentary; TPA: total physical activity.

When comparing the time in TPA from each method to the ground truth direct observation, the mean absolute differences ranged from 7.6-18.6min/hour. The ML model NVM/SED/TPA had the smallest mean absolute difference at 3.0min/hour. When comparing the time in MVPA from each method to the ground truth direct observation, the mean absolute differences ranged from 10.7-25.8min/hour. The ML model NVM/SED/LPA/MVPA had the smallest mean absolute difference at 3.2min/hour. Figure 2 shows the individual participant differences, mean absolute differences, and mean relative differences for each method for both TPA (Panel B) and MVPA (Panel C).

### Open-source model availability

The four models presented above are available with sample code here: https://github.com/LettsE/ToddlerMachineLearning. We have also developed an open-source tool (Letts Little Movers Activity Analysis©) so that the models can be used more widely, without the need for coding knowledge, shown in Figure 3 (tool details in Supplementary Material 6).

**Figure 3.**
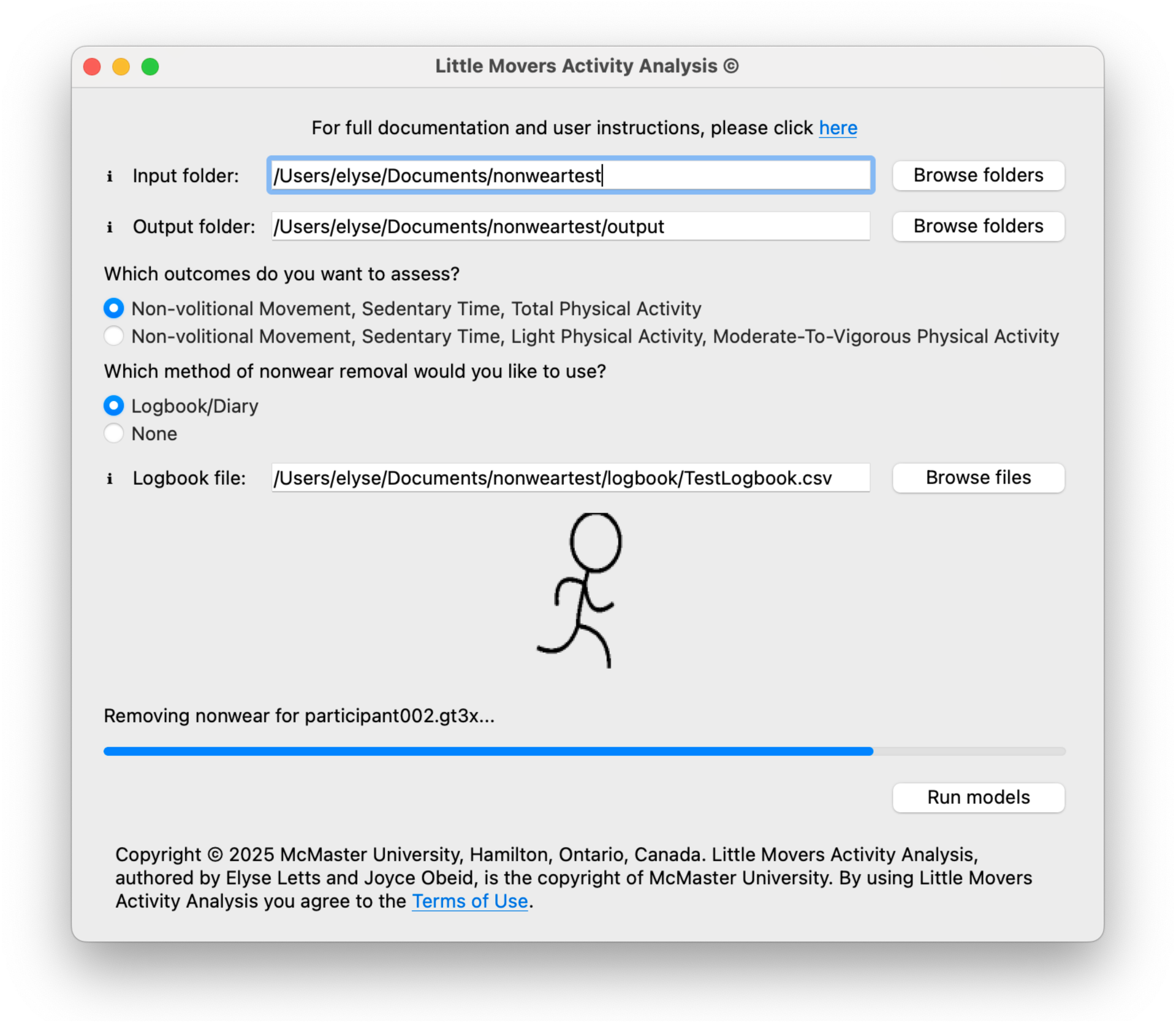
Screenshot of the Letts Little Movers Activity Analysis© tool. It provides user options for models and nonwear detection and shows the progress of the analysis.

## Discussion

This study developed open-source ML models to quantify four toddler PA and SED outcomes: (1) SED/TPA; (2) SED/LPA/MVPA; (3) NVM/SED/TPA; and (4) NVM/SED/LPA/MVPA. All models show improved accuracy over existing methods. The NVM/SED/TPA model and NVM/SED/LPA/MVPA model are recommended given their ability to filter some NVM (e.g., being carried). The NVM/SED/TPA model aligns with toddler PA guidelines while the NVM/SED/LPA/MVPA model allows for the distinction of MVPA, which may be more strongly associated with some health outcomes [2]. Our models also do not appear to overfit the data, meaning that the models are likely to generalize well to other samples. The independent sample cross-validation showed that many common methods have low accuracies and between 8-25min/hour differences in TPA and MVPA measurement.

The NVM/SED/TPA recommended model aligns with international [48] toddler PA guidelines. Its accuracy of 82% meets the generally accepted 80% threshold for model acceptability [15,49]. Its lower F1 score is reflective of the difficulty in distinguishing NVM from toddler movement, as seen in the confusion matrix with 35% of NVM misclassified as TPA. It is important to note that the 25% of NVM “misclassified” as SED, is actually correctly identified because during NVM, the toddler is sedentary. This means that the true accuracy of NVM detection is higher than the reported value. This is a substantial improvement on prior methods that have not distinguished any NVM, leading to overall greater misclassification when using cut-points. As toddlers may spend a significant amount of their day being carried or pushed in a stroller, the ability to tease out NVM is essential to avoid overestimations of PA.

Comparing activity outcomes, TPA is better classified than LPA/MVPA. This lower accuracy for LPA/MVPA is driven by LPA which is evenly misclassified as SED or MVPA. The MVPA, however, is classified quite well with an 84% accuracy. More research is needed to understand the relationships of TPA and MVPA with health outcomes using these improved activity classification models. As such, for those looking to investigate MVPA, we recommend using the NVM/SED/LPA/MVPA model.

We have made the models openly available in .json format, providing sample python code for feature extraction and model classification. We recognize that not all potential end-users are familiar with python and so have developed the Letts Little Movers Activity Analysis© tool for download. It provides a graphical user interface to batch analyse raw data in .gt3x format, optionally use a logbook file to remove nonwear time [50] (e.g., device removal, naps), then apply the model, resulting in raw classifications and summarized daily results (details in Supplementary Material 6). This free tool will help address the hurdles to ML model use [13,21].

For the secondary objective, we compared 11 published methods to the ground truth and found accuracies ranging from 33-71%. This translates to differences of 8-18min/hour of TPA and 11-26min/hour of MVPA. Even the smallest difference of 8min/hour over a 12-hour day could result in a misclassification of up to 96 minutes of TPA, more than half of the recommend amount of daily TPA. Both our NVM/SED/TPA and NVM/SED/LPA/MVPA models had a mean absolute difference of 3min/hour (using only testing data), much less than other methods.

### Clinical and research implications

Our models improve on cut-points by achieving higher accuracy and detecting some NVM – something cut-points cannot do. Compared to existing toddler ML methods, our sample is 4-10 times larger with ∼2 hours of annotated observation per participant. The combination of a large, high-resolution dataset, advanced classification, and open-source availability with an easy-to-use interface makes this a valuable tool for researchers and clinicians.

### Limitations

First, the annotation of our ground truth was completed by a relatively large number of annotators, introducing potential bias in the criterion values. To combat this, we had a rigorous training program and double-coded 10% of the videos which showed more than acceptable inter-rater metrics. Second, the recommended models have the lowest classification accuracy for NVM. While identifying any NVM is an improvement, the issue of identifying and excluding caregiver-based movement is an important avenue for future research, potentially using deep learning. Next, while our large sample allowed for a completely withheld testing set (20% of participants’ data used only for final model testing, not involved in training/tuning), which promotes the generalizability of our models to new samples, the models need validation in a fully independent sample. Finally, our participants were all healthy toddlers who were mostly White, from highly educated, English-speaking families. There is no reason to expect that these demographics would have an impact on the biomechanical aspects of movement; however, it is unclear how generalizable these models are to more diverse populations, particularly those with disabilities. These are critical avenues for future research.

## Conclusion

This study presents four novel, open-source, ML models to measure toddlers’ PA and SED. The models all had improved accuracy compared to existing cut-point methods and additionally, two of the models can detect NVM, further improving on existing methods. These recommended models assessing NVM/SED/TPA or NVM/SED/LPA/MVPA have the smallest mean absolute difference in measuring TPA and MVPA compared to existing methods by more than 5min/hour. We additionally present a free, user-friendly interface for use of these models without requiring coding knowledge. Ultimately, this study presents a substantial step forward in the measurement of toddlers’ movement.

## Data Availability

The machine learning models developed in this study are openly available along with the python code and graphical user interface: (https://github.com/LettsE/ToddlerMachineLearning). The datasets used and/or analysed during the current study are available from the corresponding author on reasonable request.

## Declarations

### Ethics approval and consent to participate

This study received ethics clearance from the Hamilton integrated Research Ethics Board (HiREB #3674). Informed written consent was given by a parent of each participant.

### Availability of data and materials

The machine learning models developed in this study are openly available along with the python code and graphical user interface (the Letts Little Movers Activity Analysis© tool): (https://github.com/LettsE/ToddlerMachineLearning). The datasets used and/or analysed during the current study are available from the corresponding author on reasonable request.

### Competing interests

The authors declare that they have no competing interests.

### Funding

This work was supported by the Canadian Institutes of Health Research (CIHR; funding reference numbers PJT-152877 and FBD-187487) and through a North American Society for Pediatric Exercise Medicine (NASPEM) Marco Cabrera Student Research Award. SKD was supported by a CIHR postdoctoral fellowship.

### Authors’ contributions

EL analysed all data, developed the machine learning models and open-source tool, and wrote the original draft of this manuscript. SKD was involved in the conceptualization, methodology development, project administration, and data collection as well as substantial manuscript editing. ND was involved in the conceptualization, methodology development, project administration, data collection, and manuscript editing. PT was involved in the conceptualization, methodology development, funding acquisition, and manuscript editing. JC was involved in the conceptualization, methodology development, funding acquisition, and manuscript editing. DK was involved in data analysis and manuscript editing. BWT was involved in the conceptualization, methodology development, project administration, funding acquisition, supervision, and manuscript editing. JO was involved in the conceptualization, methodology development, project administration, data analysis, funding acquisition, supervision, and manuscript editing. All authors reviewed and approved this version of the manuscript for submission.

## Acknowledgements

We would like to thank the participants and their families who made this research possible. We would also like to thank the Child Health & Exercise Medicine Program volunteers who assisted in the annotation of videos.

## Notes

### Competing Interest Statement

The authors have declared no competing interest.

