## Supplementary Material 1 for "Development and accuracy of a novel machine learning model to detect toddlers’ physical activity and sedentary time using accelerometers: Little Movers Activity Analysis"

**Supplementary Table 1.** Study details for existing toddler machine learning models

| Study | Number of participants | Age | Time of observation per participant | Ground truth training method | Device | Wear location | Data used to train model | ML model used | Outcomes detected | Accuracy |
| --- | --- | --- | --- | --- | --- | --- | --- | --- | --- | --- |
| Nam and Park 2013 [1] | 10 (1 for training, 10 for testing) | 16-29 months | NR | Direct observation (video annotation)  Unclear epoch/continuous | SCA3000 | Hip | Raw data features (time and frequency) extracted in a sliding ~2-3 second window | SVM | 11 activities: wiggle, roll, stand still, stand up, sit down, walk, toddle, crawl, climb up, climb down, and stop. | 86.2% (accelerometer alone), 98.4% (accelerometer + barometer) |
| Kwon et al 2019 [2] | 24 (LOSO-CV) | 13-35 months | NR | Direct observation (video annotation)  Unclear epoch/continuous | ActiGraph wGT3X-BT | Hip, wrist | NR but 2 second window | Random forest classifier (also tested SVM, decision tree, K-nearest neighbors, logistic regression) | 8 activities: walk/run, climb up/down, stand, crawl, sit, lie down, carried, and riding a stroller/wagon | 69% (hip), 55% (wrist) |
| Kwon et al 2019 [3] | 21 (LOSO-CV) | 13-35 months | 8-25 minutes | Direct observation (video annotation)  Continuous annotation | ActiGraph wGT3X-BT | Hip | Sum of activity counts in x, y, z, and vector magnitude in 5s windows | Random forest classifier | Carried vs ambulation | 89% |
| Albert et al 2020 [4] | 22 (LOSO-CV) | 13-35 months | 6-21 minutes | Direct observation (video annotation)  1s annotation | ActiGraph wGT3X-BT | Hip | 76 raw data features (time and frequency) in a 2s window | Random forest classifier with Hidden Markov Model augmentation | 8 activities: run/walk, crawl, climb, stand, sit, lie down, carried, and stroller | 64.8% |
| Thornton et al 2023 [5] | 279 | 9-38 months | NA | NA: unsupervised machine learning benchmarked to cut-points | ActiGraph GT3X+ | Hip | Mean ENMO in a 10s window | Hidden semi-Markov model | 3 intensities: sedentary, light PA, moderate-to-vigorous PA | NA |

*Note.* ENMO: Euclidean Norm Minus One; LOSO-CV: leave-one-subject-out cross validation; NA: not applicable; NR: not reported; PA: physical activity; SVM: Support Vector Magnitude
