## Supplementary Material 2 for "Development and accuracy of a novel machine learning model to detect toddlers’ physical activity and sedentary time using accelerometers: Little Movers Activity Analysis"

### iPLAY Coding Definitions

**Each intensity and position code is defined as follows:**

**Intensity codes for movement behaviour**

**1 -** stationary, no movement of ANY limbs
- hands/feet moving with no limb movement
- Slight head rotation allowed with no trunk movement
- must be completely stationary for the **whole second duration**

**2 -** stationary & moving limbs and/or minimal trunk displacement
- moving from elbow/shoulder or knee/hip
- slight shifting weight
- leaning while stationary (minor/**barely noticeable** trunk displacement)
- minor sway

**3** –full trunk moving – intention to move trunk/**obvious** trunk movement
- twisting, reaching (obvious truck displacement)
- up and down (e.g. sit to stand, stand to squat)
- if the trunk is moving but there is no **intent** to move from one location to another this would be coded as 3 (stepping in place, stationary pivoting – moving one foot while other foot remains in the same place)
- stationary turning around (on the spot)

**4** – SLOW/USUAL translocation of the entire body (moving through space)
- their **intent** must be to move from one place to another
- lateral, forward, backward - stepping either foot in any direction with intent to move body to another location
- turning/changing direction as part of continuous movement to another location
- Jumping can be coded as a 4 or 5 depending on how intense/fast/hard the jump is. Example:
  - Hesitant, cautious, slow jump = 4
  - Jumping over small hurdle/standing long jump = 4

**5** – FAST translocation of the entire body (moving through space)
- this is relative for the child when they are moving FASTer than normal/usual pace
- their **intent** must be to move from one place to another
- Jumping can be coded as a 4 or 5 depending on how intense/fast/hard the jump is. Example:
  - Big, fast jump off the bigger stairs = 5
  - Jumping over big hurdle/ running jump = 5

**6 -** being carried or moved not of their own volition (e.g., parent/examiner moving their limbs, parent is carrying child while walking)
- if parent could be replaced with an inanimate object (e.g., chair, handrail, table), then code child's movement (e.g., if parent is stationery and child is sitting/standing/climbing on parent then code child's movement).
- if parent is using hands to help child walk/climb stairs the child's movement should be coded (do NOT code 6)
- if unsure whether the movement is child's own volitional movement or initiated by parent code 6

**7 -** being pushed/pulled (e.g., stroller, wagon)

**8** – pulling, tugging, touching accelerometer
- be liberal, even if possibly touching, moving accel and/or shirt code as 8

**9** – Accelerometer not being worn

**0** - NOT IN PICTURE.

- Code 0 if any limb is out of view when you are unsure whether it should be a 1 or 2
    - If a portion of the child is out of view but the code would stay the same regardless of what the missing portion is doing you can still code appropriately (For example: child is walking/running and their arm is out of view you can still code appropriately, but if a child is sitting still and you can't see what their arm is doing you wouldn't be able to code (could be a 1 or a 2)
- Never assume what is happening when the child is out of view, code 0 if unsure or if out of view at any point during the second epoch

**X** – Falling (e.g. running and trip, lose their balance while standing and fall to floor)

**Additional Notes**

- In some instances, to understand the child's **intent** play the video at full speed
    - Child is going up/downstairs should be coded 4, even if very slow, unless clear pause in movement up or down when watching in real time

- The greatest intensity movement (1-5) in that second is coded (i.e., 2 trumps 1, 5 trumps 4), even if only occurs briefly within the second epoch

- If 6,7,8, 9, 0 occurs anywhere in the epoch this is coded instead of the intensity (1-5)

- Play it safe, if unsure code 6,7,8,9 or 0 instead of an intensity category, can flag in the Notes

- If unsure of what to code, code what you think is best and leave a detailed note explaining what you are unsure about

**Position codes**

**A** – Sitting position on ground (sitting, squatting, crawling, reclining in chair, scooting)
**B** – Standing position
**C** – Lying position (body is horizontal, back or trunk/chest must be on ground, even partially)
**O** – Other, for when 6,7,8,9, or 0 are coded for intensity

- When participation transitions between different positions code starting position then ending position (ie. AB for sitting to standing; AC for sitting to lying, etc.)
- Must be in position for the **whole second duration**, or else code transition
