## Supplementary Material 3 for "Development and accuracy of a novel machine learning model to detect toddlers’ physical activity and sedentary time using accelerometers: Little Movers Activity Analysis"

**Supplementary Material 3 – Results of epoch analyses**

**1 second epoch**

Accuracy: 0.72

F1: 0.58

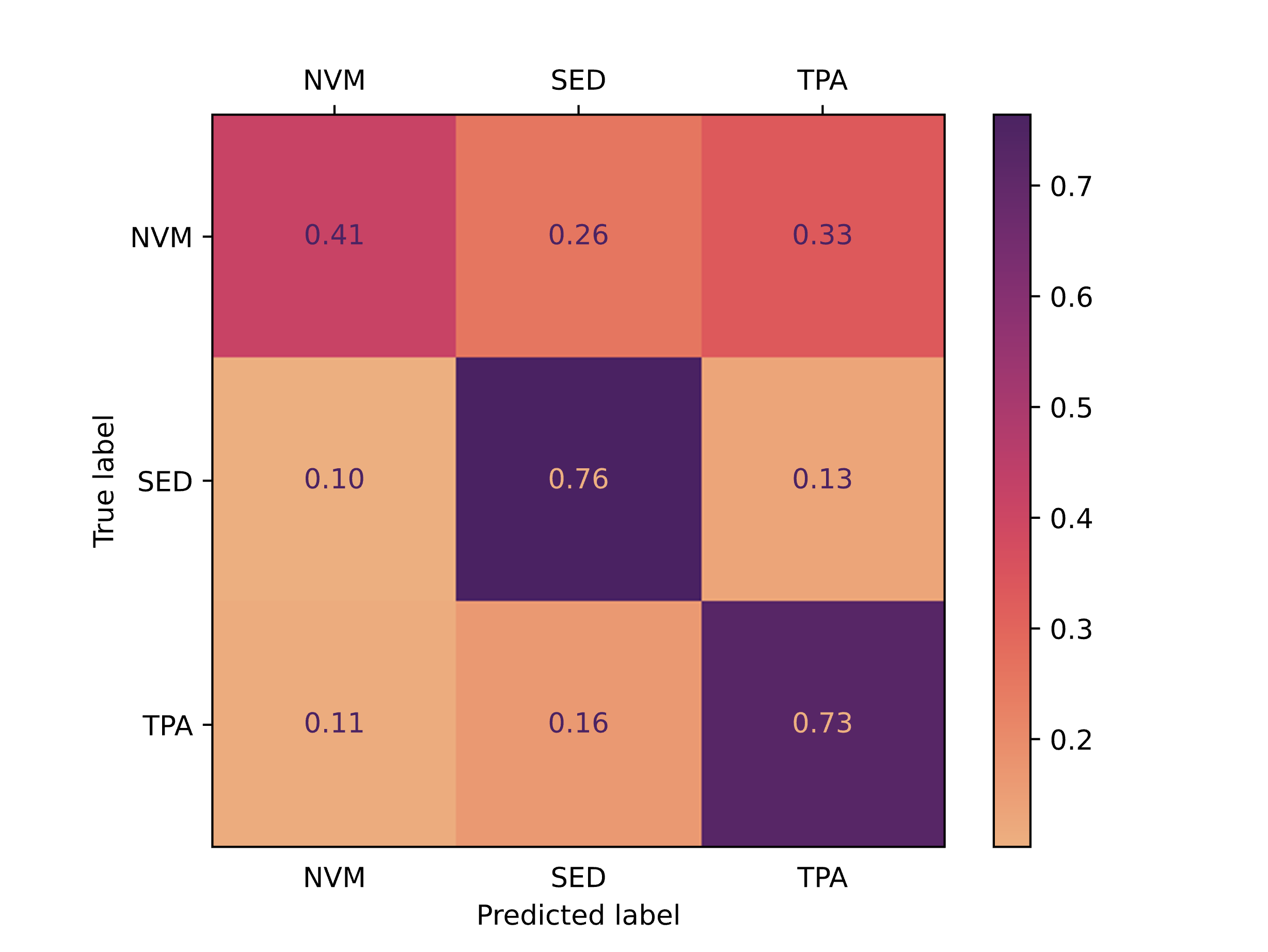

**3 second epoch**

Accuracy: 0.80

F1: 0.65

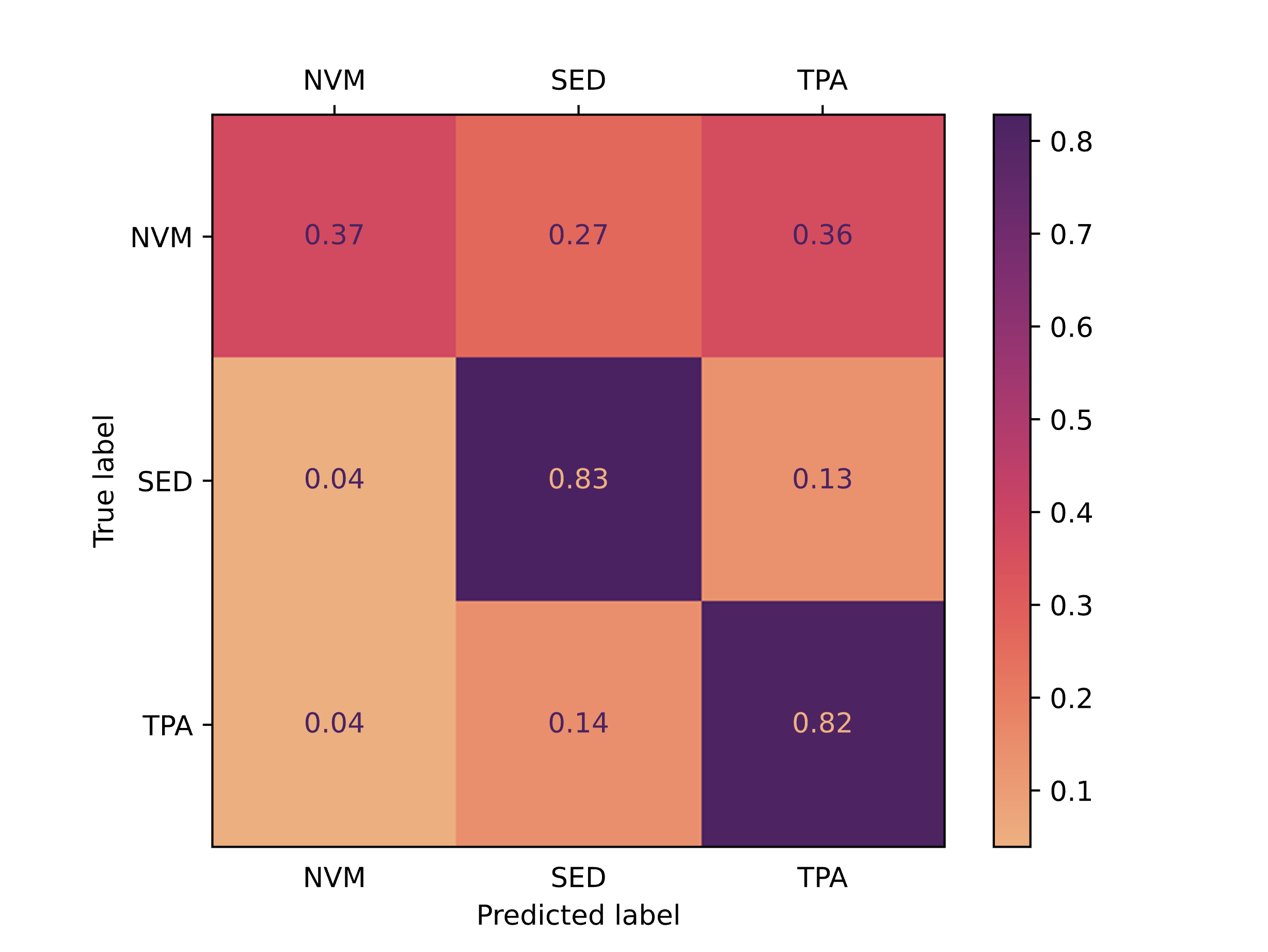

**5 second epoch**

Accuracy: 0.82

F1: 0.69

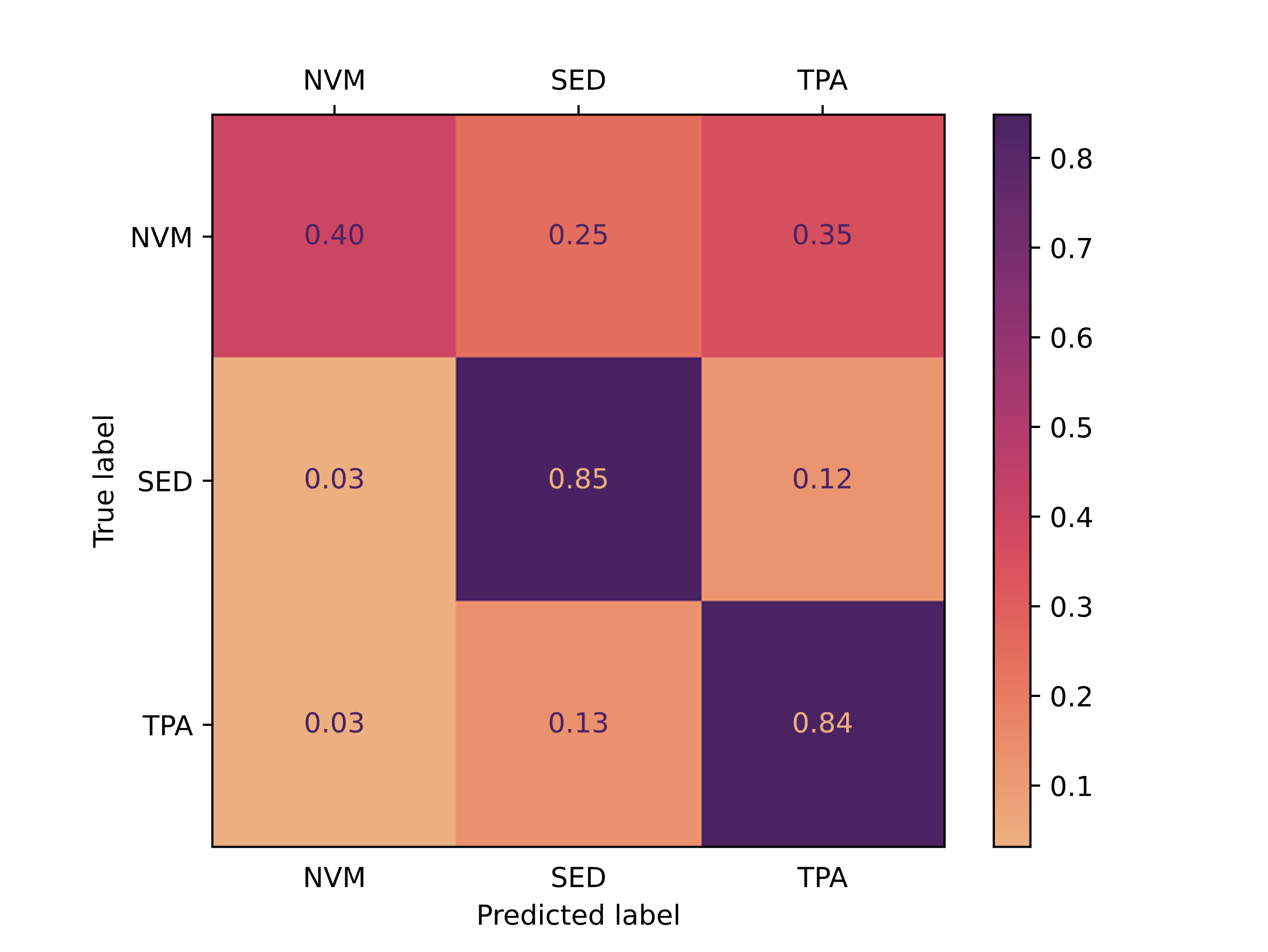

**15 second epoch**

Accuracy: 0.83

F1: 0.69

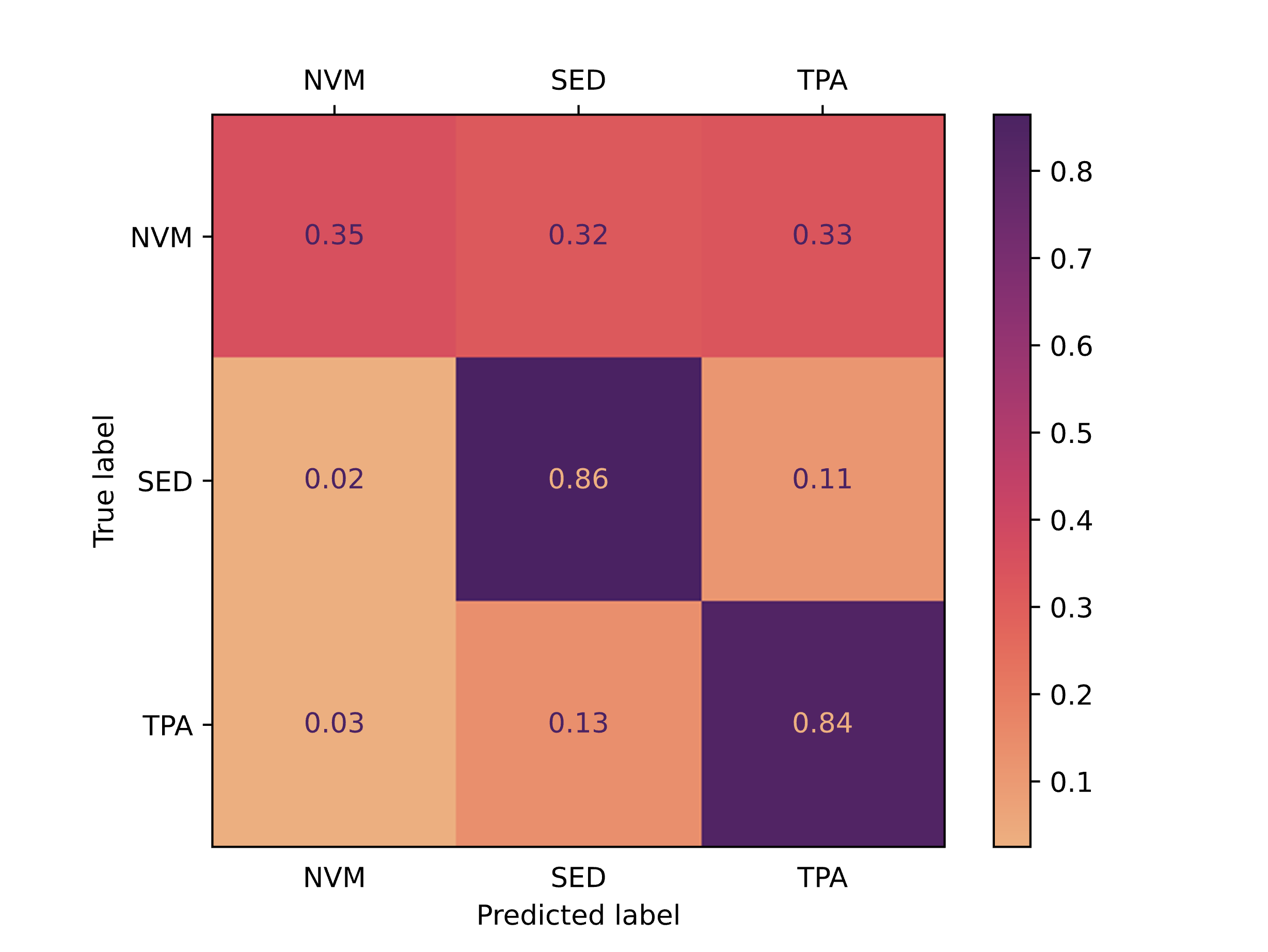

**30 second epoch**

Accuracy: 0.83

F1: 0.68

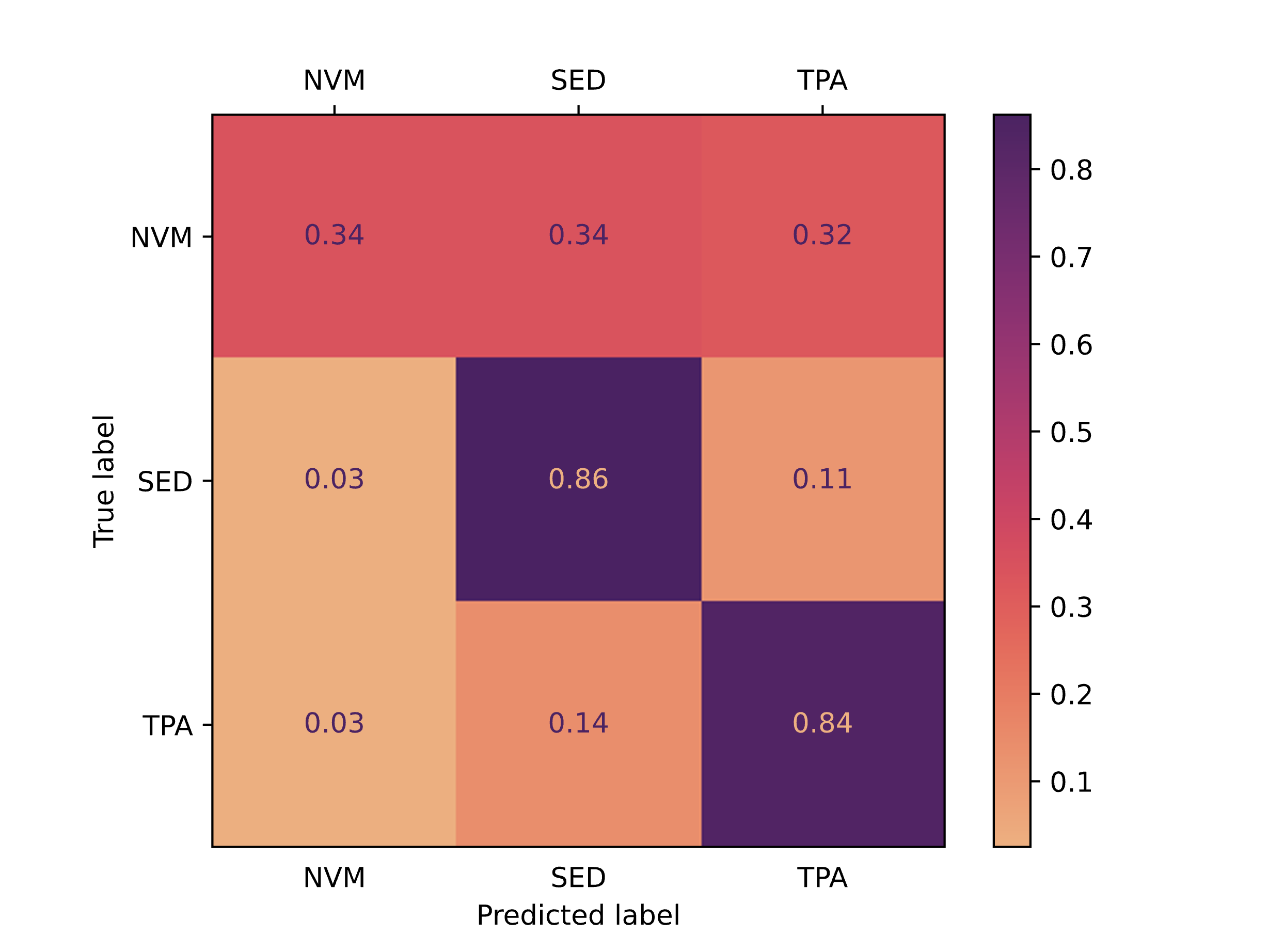

**60 second epoch**

Accuracy: 0.82

F1: 0.66

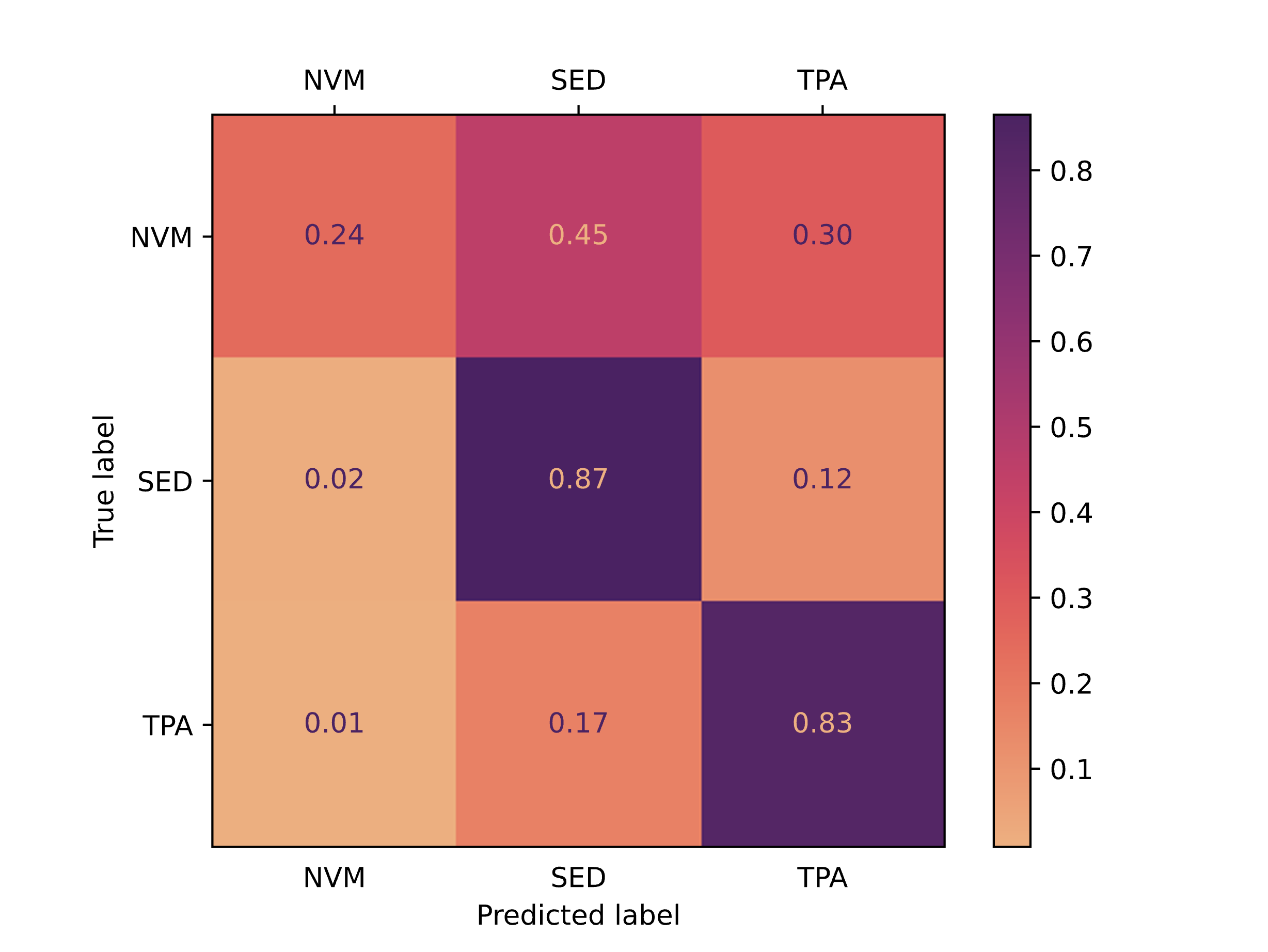
