## Supplementary Material 4 for "Development and accuracy of a novel machine learning model to detect toddlers’ physical activity and sedentary time using accelerometers: Little Movers Activity Analysis"

**Table 4**. Frequency of video annotation observations in 5s epoch.

| Code | Frequency |  | Category | Frequency |
| --- | --- | --- | --- | --- |
| 1 | 12868 |  | SED | 53257 |
| 2_other | 40389 |  |  |  |
| 2_standing | 16014 |  | LPA | 38610 |
| 3 | 22596 |  | MVPA | 46837 |
| 4 | 44031 |  | TPA (sum of LPA and MVPA) | 85447 |
| 5 | 2806 |  |  |  |
| 6 | 6229 |  | NVM | 6637 |
| 7 | 408 |  |  |  |
| 8 | 3866 |  | Invalid (8, 9, X, 0) | 15025 |
| 9 | 11146 |  |  |  |
| X | 13 |  |  |  |
| 0 | 6544 |  |  |  |
