## Supplementary Material 6 for "Development and accuracy of a novel machine learning model to detect toddlers’ physical activity and sedentary time using accelerometers: Little Movers Activity Analysis"

**Little Movers Activity Analysis Overview**

The Letts Little Movers Activity Analysis tool is a graphical user interface that allows for the estimation of toddler sedentary time and physical activity using machine learning models. Full instructions for downloading and using the most up-to-date version of the tool are available on the GitHub page: <https://github.com/LettsE/ToddlerMachineLearning>

**How do I download it?**

To download the Letts Little Movers Activity Analysis tool, please follow these steps:

1. From the GitHub page, the most up-to-date version is located in the "Releases" section (right side panel).
2. Download the zip file for your operating system (Windows, macOS).
3. Once downloaded, unzip and open the file.
4. Double click on "LittleMoversActivityAnalysis.app" (macOS) or "LittleMoversActivityAnalysis.exe" (Windows) to open the tool.

**How do I use it?**

Once you have downloaded and opened the tool, you are ready to run your data:

1. Choose your input folder: Select "Browse" to open the file selector and choose the folder where your gt3x files are located. .gt3x files should be named using the studyid/participant id (e.g., participant001.gt3x)
2. Choose your output folder: Select "Browse" to open the file selector and choose the folder where you want the output files to be saved.
3. Choose model outcomes: select between two models. The first option has outcomes of non-volitional movement, sedentary time, and total physical activity. The second option has outcomes of non-volitional movement, sedentary time, light physical activity, and moderate-to-vigorous physical activity. See paper for recommendations on which model to use.
4. Choose nonwear method: Current options include "None" so all data will be passed to the model or "Logbook/Diary" which will removed nonwear times listed in the logbook.
5. (Optional) If Logbook/Diary is selected for nonwear, select "Browse" to open the file selector and choose the .csv file that contains the logbook/diary information. This .csv file must include headers studyid, WearTimeStart, and WearTimeEnd. WearTimeStart/End should be in Datetime format: 2025-03-16 07:14:54
6. Click on Run models. A progress bar will appear to track progress of all files in the input folder. If you have missed a selection, it will prompt you to finish the selections before running the models.

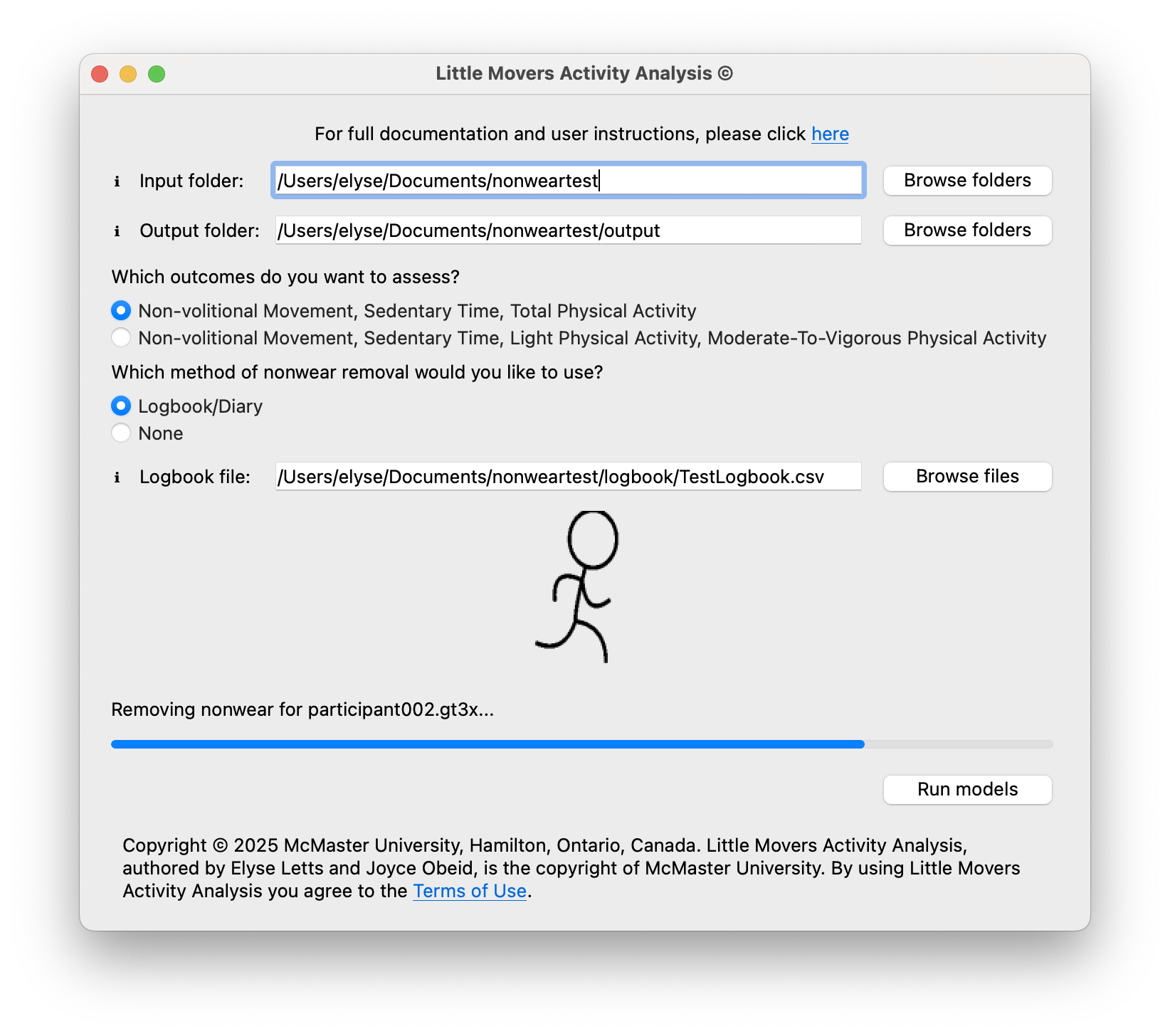

**How many files can I run at once?**

While there is no limit to the number of files that can be in the input folder, we recommend running a **maximum of 10 .gt3x files** in each instance of the Letts Little Movers Activity Analysis tool. This is to avoid overwhelming your computer or crashing the tool while running. To run more than 10 files at a time, we recommend opening more than one instance of the Letts Little Movers Activity Analysis tool (double click on "LittleMoversActivityAnalysis.app/.exe" to open another instance). For 7 days of accelerometer wear, it takes ~25 minutes to process one file.

**What are the output files?**

There are 6 possible output files (depending on user selections) generated:

Always generated:

1. {filename}_predictions.csv (e.g., participant001.gt3x_predictions.csv): This csv contains the epoch-by-epoch features and final model prediction for an input .gt3x file. The first 160 columns contain the features used by the model followed by the column "Prediction" (contains one of "NVM", "SED", "TPA", "LPA", or "MVPA"), then Time (Datetime timestamp). This output is likely most helpful for individuals wishing to run an independent sample cross-validation of the models or who need a very high level of granularity to the data.
2. by_day_by_participant.csv: This contains the time (in minutes) in each of the model outcomes (NVM/SED/LPA/MVPA/TPA) for each participant each day. If no logbook/diary was used to remove nonwear, this will be the final file that can be used for further analysis. When completing further analysis, we recommend summing NVM and SED to obtain the total sedentary time of the toddler.

Only generated when a logbook/diary is used to remove nonwear time:

3. {filename}_trimmed_data.csv (e.g., participant001_trimmed_data.csv): It contains the raw data (X, Y, Z) with vector magnitude and timestamp for only the wear time. This file can be used to check that nonwear time was removed properly.
4. wear_daily_summary.csv: This contains the wear time (and corresponding nonwear time) for each participant (studyid) and day (Date).
5. all_wear_time.csv: This contains the final wear times that were used to trim the data. If all data was correctly formatted, this should be a duplicate of your input logbook/diary file. It can be used to help troubleshoot any discrepancies in nonwear removal.
6. FinalSummaryByParticipant.csv: This combines the wear_daily_summary and by_day_by_participant files into one summary .csv file that can be used for further analysis. When completing further analysis, we recommend summing NVM and SED to obtain the total sedentary time of the toddler.

**Can anyone use the models and the Letts Little Movers Activity Analysis tool?**

Yes, anyone can use both the models and the Letts Little Movers Activity Analysis tool provided that they agree to and follow the copyright and Terms of Use (<https://github.com/LettsE/ToddlerMachineLearning/blob/main/LICENSE>) and that they cite the paper associated with the models.
